# Modeling spatiotemporal *Aedes aegypti* risk in French Guiana using meteorological and remote sensing data

**DOI:** 10.1101/2021.08.02.21261373

**Authors:** Sarah Bailly, Vanessa Machault, Samuel Beneteau, Philippe Palany, Romain Girod, Jean-Pierre Lacaux, Philippe Quenel, Claude Flamand

**Author notes:** University of Bordeaux, Bordeaux, France (Vanessa Machault).

## Abstract

Although the development of vaccines for the prevention of arboviral diseases has been a priority in recent years, prevention strategies continue to depend on vector control. Risk maps at scales appropriate for these strategies can provide valuable information to assess entomological risk levels and guide actions. We used a spatio-temporal modeling approach to predict, at the local scale, the risk of homes potentially harboring *Aedes aegypti* larvae. The model used integrated larvae risk data collected in the field from September 2011 to February 2013, environmental data obtained from very high spatial resolution Pleiades imagery, and daily meteorological data, collected in the city of Matoury in French Guiana. Various environmental and meteorological conditions were identified as risk or protective factors for the presence of immature stages of *Aedes aegypti* in homes on a given date and used to produce dynamic maps with high spatial and temporal resolution. Aedes vector risk was modeled between 50 and 200 m, around houses, on a time scale of 3 to 5 days. The resulting model was extrapolated to other municipalities with the same characteristics of urbanization during the 2019-2020 dengue epidemic in French Guiana. This work represents a major opportunity to monitor the evolution of vector risk and constitutes information that could be particularly useful for public health authorities in charge of vector control.

## Introduction

Arboviral diseases are caused by viruses that are transmitted to humans through the bite of an infected arthropod. Although arboviruses have been infecting humans for a long time, in recent decades they have become a growing public health problem due to the emergence and re-emergence of the diseases they cause throughout the world (Gould *et al*., 2017, Gould *et al*., 2010, Petterson *et al*., 2016, Bhatt *et al*., 2013, Kraemer *et al*., 2019, Gubler, 2011). In Latin American and Caribbean countries, the reintroduction and dissemination of *Ae. aegypti* took place in the 1970s (Brathwaite *et al*., 2012), leading to a progressive increase in the risk of arboviruses and the regular occurrence of large-scale epidemics (Brathwaite *et al*., 2012, Halstead, 2006, Martin *et al*., 2010).

In French Guiana, an overseas French department of 300,000 inhabitants located in the northeast region of the South American continent along the Atlantic coast, several epidemics linked to arboviruses have occurred in recent decades. Between 1991 and 2013, large-scale dengue epidemics occurred in a 3-4 years cycle (Adde *et al*., 2016, L’Azou *et al*., 2014, Flamand *et al*., 2014). Between 2006 and 2013, four dengue epidemics associated with one or two serotypes occurred in succession, each of them causing between 9,000 and 16,000 clinically dengue-like syndroms. During the 2013 epidemic, 16,000 clinical cases were recorded in this region with 689 hospitalizations and 6 deaths (Flamand *et al*., 2017). Following their recent introduction in the Americas, the chikungunya and Zika viruses caused two major epidemics in 2014 (Flamand *et al*., 2019, Fritzell *et al*., 2018, Flamand *et al*., 2017). After a period during which there was no dengue virus circulation in the department between 2015 and 2018, dengue virus circulation gradually intensified until a new epidemic started in 2020 (Santé Publique France, 2020), related to two serotypes and generating 12,300 clinical cases and 4 deaths. (Santé Publique France, 2021).

The *Aedes aegypti* mosquito is the only identified vector for the transmission of dengue, chikungunya and zika viruses, in French Guiana. The mosquito reproduces mainly in domestic containers of clean water with little organic debris. Plant containers, abandoned detritus, discarded cars and pneumatic tyres, poorly maintained gutters, old household appliances and swimming pools are often filled with rainwater and represent potential breeding sites.

Although the development of vaccines for the prevention of arboviral diseases has been a priority in recent years, prevention strategies continue to depend on vector control. In French Guiana, viral circulation control relies on indoor and outdoor deltamethrin spraying to control adults mosquitoes adult mosquitoes in homes where human cases have been identified and in homes within a 100 m radius. Larval breeding sites are eliminated mechanically or chemically. In addition to these vector elimination actions, a health education program based on home, to neighboring homes and school visits has been deployed (Epelboin *et al*., 2018, Dusfour *et al*., 2015). Despite this, the effectiveness of these actions is reduced by the lack of coverage of cases reported to the health authorities as well as the sometimes long delays, due to the sometimes approximate location of contamination sites, between the biological confirmation of cases and the intervention of control operators. Combined with epidemiological data, entomological data that can inform about vector risks, collected at the local level on a repeated basis would be necessary to understand and intervene rapidly in the areas where the transmission risk is the most important.

Unfortunately, these data are often collected in a discontinuous manner and provide imprecise information to effectively identify areas to prioritize for control actions. In order to establish control strategies, fine-scale risk maps are needed to know precisely the areas of high probability of transmission due to the presence of larval breeding sites. Knowing at a given time, the exact locations associated with high larval transmission risk would allow targeting interventions and optimizing the control efforts of vector control actors.

Weather and climate conditions (rainfall, relative humidity, and temperature) (Barbazan *et al*., 2002, Otero *et al*., 2010) as well as environmental conditions (vegetation, soil type, urbanisation rate) (Eisen and Lozano-Fuentes, 2009) play important roles in the evolution of dengue epidemics. These conditions are spatially and temporally distributed in very heterogeneous patterns. As a useful tool for collecting environmental and meteorological data with the choice of spatial, spectral and temporal resolution, remote sensing is used to monitor these conditions at global, regional and local scales.. This tool was combined with different machine learning algorithms to detect related associations with entomological indices. In 2014 in Martinique, Machault *et al*. used logistic regression to model the presence of houses potentially hosting *Aedes aegypti* larvae. The presence of larva was significantly associated with predictors of maximum humidity and asphalt surface. In Thailand, Sarfraz *et al*. used a decision tree to predict the probability of vector reproductive habitats using temperature, rainfall, human density, land cover and elevation indicators. Others researchers have used time series (Lana *et al*., 2014) or spatial analysis (De Melo *et al*., 2012) to map abundances and investigate correlations with reported cases. The results of these studies have suggested that the effects of climatic parameters on the incidence of dengue epidemics can vary considerably from one study site to another (Liao *et al*., 2015, Ferreira, 2014) and are largely dependent on the local epidemiological context models. There is currently no model in French Guiana to help develop an early warning system for entomological risk. This article therefore aims to understand and model the fine spatial (house) and temporal (daily) dynamics of the presence/absence of *Aedes aegypti* larvae according to meteorological and environmental factors, in a study area located in Matoury, a municipality on the French Guiana coast in the vicinity of Cayenne. This larval model was then extrapolated to all houses in the municipalities surrounding Cayenne including Cayenne, Rémire-Montjoly and Matoury municipalities during the last dengue epidemic in order to evaluate the capacity of our model to target high transmission risks in other municipalities.

## Results

### Model results

Larval data were available for 261 different houses surveyed between 1 and 3 times, comprising a total of 333 observations, of which 41% were positive for the presence of *Aedes* aegypti larvae. The final BRT model was reduced from 317 variables in the logistic regression to 6 explanatory variables containing 3 environmental variables corresponding to 59% of the cumulative influence of the model and 3 weather variables corresponding to 41% of the cumulative influence (Table 1). The variable with the highest response was the low vegetated soil surface in a 100 m zone around each house. Its relative positive influence was 26%. The average temperature in the previous 5 days and the average Redness Index (RI) within 50 m around each house and the average normalized difference turbidity index (NDTI) in a 200 m buffer zone had a relative importance value of 19%, 18% and 15% respectively. The two model variables with the lowest relative influence were the average minimum temperatures in the previous 25 days and the cumulative precipitation in the previous 3 days. These variables had relative importance values of 12% and 10%, respectively (Figure 1).

**Table 1:** Associated factors.

|  | **Name of the variable** | **Relative importance** |
| --- | --- | --- |
| Surf_low_veget_100m | Surface of low vegetation_100m | 26% |
| Mean_temp_5_days | Mean temperature in the 5 last days | 19% |
| Mean_RI_50m | Mean RI_50 meters | 18% |
| Mean_NDTI_200m | Mean NDTI_ 200 meters | 15% |
| Min_temp_25_days | Minimal temperature in the 25 last days | 12% |
| Rainfall_3_days | Rainfall in the 3 last days | 10% |

**Figure 1:**
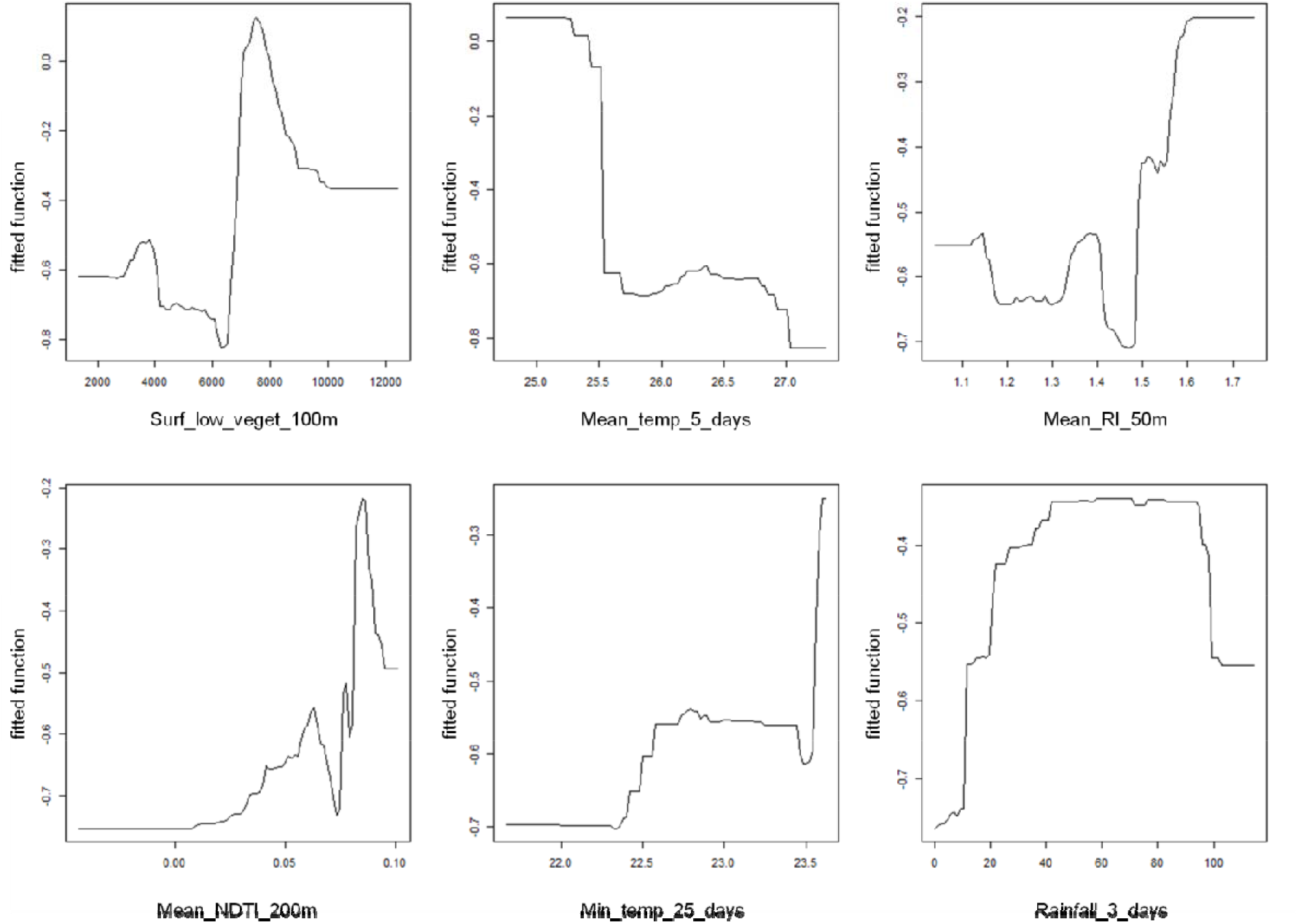
Graphs of associations between explanatory variables and the presence of larvae.

Figure 1 shows the directions of the associations and thresholds at which the predictions changes. It can be seen that at a mean temperature of 25.5°C in the previous 5 years, the probability of the presence of *Aedes aegypti* larvae decreases. For the RI, the tendency is the opposite. From the cutoff point of 1.5, the probability of the presence of larvae increases. Finally, the cumulative rainfall in the previous 3 days constitutes a high probability plateau of the presence of *Aedes aegypti* larvae, between 10 mm and 100 mm.

An entomological situation variable was added to the 6 other variables to improve the prediction of the risk in a given house on a given date. This variable corresponds to the average of the probabilities of presence of larvae in the houses present in a radius of 30 m and over the previous 7 days and was created using the first run of the model described above and then fed into the second modeling “run” used to predict the presence/absence of *Aedes* larvae for each geolocated house. The area under the ROC curve was 0.72.

### Larvae risk mapping

During the 10 months of the studied epidemic period of 2019-2020 and for each house, the extrapolated model (Figure 2) shows whether there is a risk of presence (red) or absence (blue) of *Aedes* Aegypti larvae. No house is at risk on every day of the study period.

**Figure 2:**
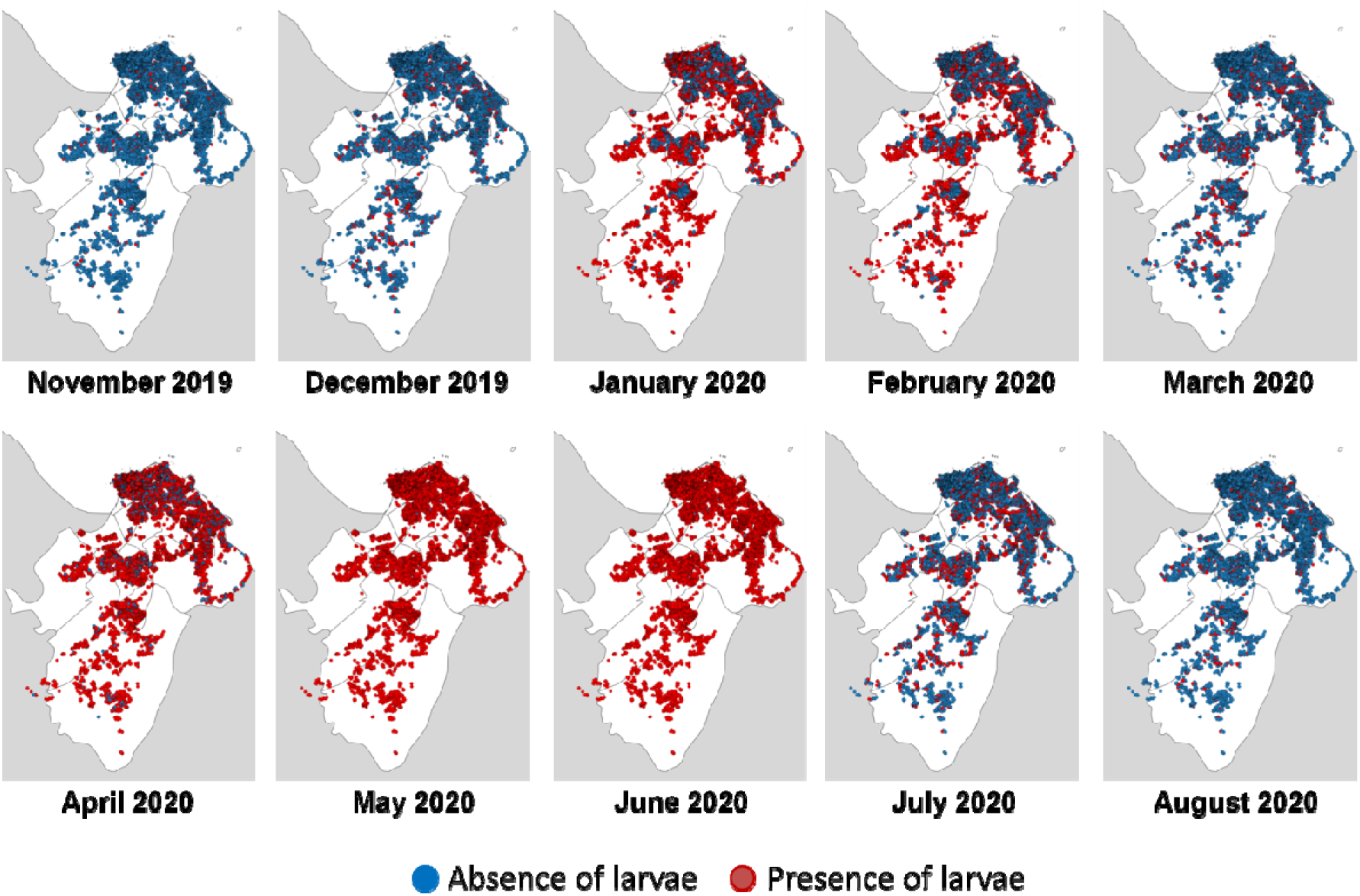
Monthly entomological risk maps from the modeling experiment based on data from November 2019 until August 2020 in theCayenne municipality.

Similarly, all buildings appeared to be at risk for at least 100 days. The positivity rate varied between 3.8% and 99.6% (Table 2). The months with the highest entomological risk were May and June. During these two months, the positivity rate for the presence of larvae was above 99%. On the other hand, the months at the beginning and end of the epidemic period were those in which the positivity rate was lowest. Thus, in November the rate is 3.8%. For the month of August, the rate of positive larvae presence was 6.6%.

**Table 2:** Correlation rate between entomological and epidemiological data.

|  | **Number of positive cases** | **Number of positive houses** | **Rate of positive houses (%)** | **Correlation rate** |
| --- | --- | --- | --- | --- |
| November 2019 | 3 | 1125 | 3,8 | 0.2 |
| December 2019 | 0 | 2339 | 7,9 | - |
| January 2020 | 8 | 15796 | 58,6 | 0.3 |
| February 2020 | 11 | 10730 | 38,6 | 0.2 |
| March 2020 | 29 | 3707 | 9,0 | 0.2 |
| April 2020 | 79 | 20874 | 72,8 | 0.3 |
| May 2020 | 121 | 25610 | 99,6 | 0.4 |
| June 2020 | 156 | 25608 | 99,6 | 0.6 |
| July 2020 | 168 | 6890 | 24,6 | 0.5 |
| August 2020 | 122 | 2033 | 6,6 | 0.5 |

The highest positive correlations between the entomological and epidemiological variables were found with a 1000 m x 1000 m grid. Correlations were highest between June and August 2020 (Table 2) Choropleth maps (Figure 3), showed spatial heterogeneity of entomological and epidemiological results. In November and December 2019, at the beginning of the epidemic, the number of cases was low, the number of positive houses was low and no particular pattern emerged. As the number of cases increased and the number of predicted positive houses increased, an increasing number of tendencies appeared (darker grid). In April, May and June some grids appeared as hotspot areas.

**Figure 3:**
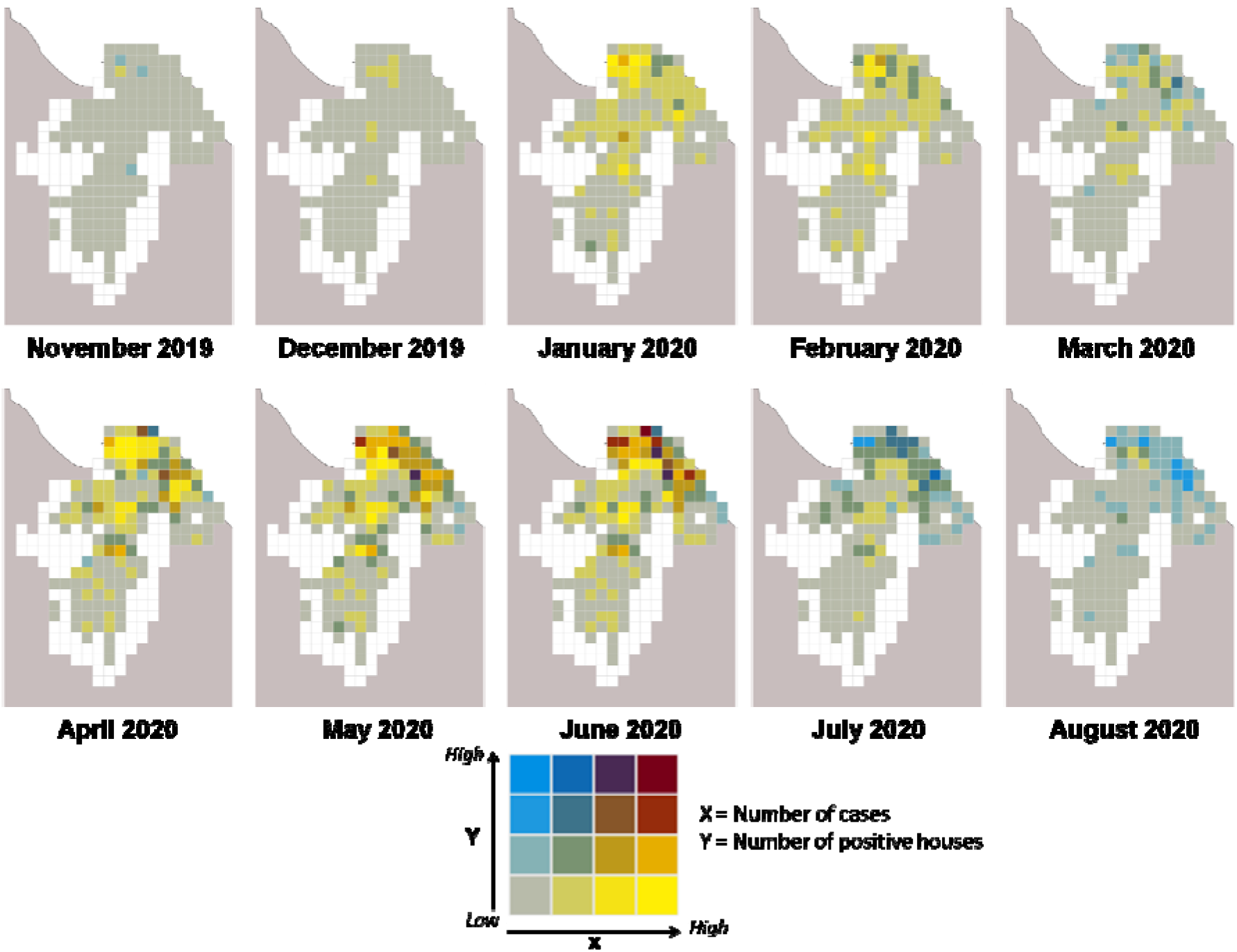
Correlation between the number of cases and the number of positive houses for the months where the model was extrapolated.

## Discussion

In this study we investigated the climatic and environmental factors of the *aedes* mosquito, the main vector of arboviruses in French Guiana, in order to predict spatiotemporal dynamics of entomological risk of larvae presence/absence. Environmental information collected using very high-resolution satellite imagery coupled with daily meteorological field data was successfully highlighted as determinants for the presence of *Aedes aegypti* larvae.

The environmental variables represented the surroundings of the surveyed houses and the spatial variability in the model as they depended on the geographical position of each house. Close and more distant environment of the houses were investigated considering 50, 100 and 200 m radius area, No larger scale variables were associated with larvae risk, indicating that the biological and physical phenomena leading to the presence of larvae occur rather close to houses. For the lightly vegetated soil of the “wet” season in a 100 m radius area, the probability of larvae presence increased with the surface area, with a cutoff effect observed for a median surface. This effect has already been demonstrated in Martinique, where the surface area of poorly vegetated soil was found to be a risk factor for the presence of water lodges (which was not necessarily positive for larvae), at approximately the same scale (Machault *et al*., 2014). These water lodges were interpreted as resulting from a lack of maintenance of the surroundings of houses, potentially generating the presence of these deposits. Our findings showed that the soil type influences the presence of mosquitoes within a 50 m radius. The characteristics of the surroundings of a house (yard, garden) impact the presence or absence of water deposits and the ability of larvae to develop in their deposits. Correlations of RI with vegetation and water-related indices to aid interpretation have been sought unsuccessfully. Soil characteristics have a relationship with known vegetation characteristics. Thus, depending on the soil and the vegetation on it, the presence of *Aedes aegypti* may be favored by providing shade on the roost (Vezzani and Albicocco, 2009). Moist soils at low temperatures favor larval development (Hemme *et al*., 2009). Finally, the supply of nutrients in the water at the breeding sites (Reiskind *et al*., 2009), attracts females for egg-laying and allows larvae to feed; well-nourished larvae develop better and have better survival rates (Tun-Lin *et al*., 2000). They also generate larger females which in turn increased the mosquito’s survival. A high density of larvae in a location is evidently partly related to the presence of a high density of females. The NDTI in a 200 m buffer indicates bare rooftops and soil, i.e. urbanization. This index plays a positive role with a threshold. Urbanization provides them with artificial lodgings and blood meals, at this 200 m scale.

We have also identified three meteorological variables that are static in space and represent both the situation on previous days and the seasonal level. The average of the mean temperatures in the previous 5 days shows that the probability of larvae presence as high as 25.5°C. This effect is in line with the above mentioned effect of the water temperature of the breeding sites which is unfavorable to larvae when growing. In addition, high air temperatures are not conducive to adult survival. The average minimum temperature in the previous 25 days was positively associated with the presence of larvae with 2 thresholds at ∼22.5 and 23.5°C. Cool seasons are therefore not conducive to the presence of vectors. Finally, the cumulative rainfall in the previous 3 days was positive and quickly reached a limit. When it rained more than ten millimeters in three days, the probability of the presence of larvae reached its maximum. In the study area, the breeding sites were small (cups, pots, and watering cans) and were therefore quickly filled with water. The extrapolation of the model showed that March was less favorable for the presence of larvae than the other months of the rainy season. This month corresponds to the “short March summer”, when rainfall decreases significantly, between the short rainy season (January-February) and the long rainy season (April-June). As soon as the main rainy season starts, the probability of larval presence increases again.

The spatiotemporal risk maps produced in this study represent an important contribution to the development of targeted operational control systems for arboviral diseases in urban areas.

It is largely accepted that entomological risk alone is not totally predictive of the occurrence of human cases of arbovirus, as other factors are also important (socioeconomic level, virus circulation, means of protection against bites, etc.). However, the presence of vectors is a particularly important element in assessing the risk of transmission, especially when associated with the circulation of viruses. The proposed approach aims to identify, from the chloropleth maps, areas at high risk of transmission due to the combined presence of the vector risk and the presence of the virus. These maps can help direct vector control actions to hotspots areas. In addition, limitations may exist in collected entomological data. For larvae, the productivity of the breeding sites was not considered and it might have been interesting to model only the presence of larvae at the most productive sites. However, it is not certain that this is a problem in the specific case of French Guiana as productivity was fairly dichotomous: the productivity was close to 20% at positive sites with buckets/watering can containers and plants, versus close to 0% for all other types of sites. Another limitation of the study is based on the methodology used to explore the associations between modeled entomological risk and cases occurrence. Indeed, the cases included in our study were determined from the surveillance system for biologically confirmed cases. Although this surveillance system is effective in the early detection and monitoring of epidemic dynamics, it does not allow for the exhaustive detection of all dengue cases. Consequently, the number of actual cases is higher than the number of cases recorded by the surveillance system, as several cases of dengue may have occurred in the control houses.

In the medium term, it will be interesting to identify geo-localized indicators that are more representative of epidemiological risk than individual confirmed cases to better highlight the link between entomological risk and epidemiological risk. The next steps will include new strategies to produce such high resolution local maps throughout the country in order to intensify entomological surveillance while reducing labor costs. It will be important in the future to set up collaborative work with control operators to appropriate this type of tool to improve research strategies.

## Materials and methods

### Study area

French Guiana is located in South America (Figure 4), north of the equator, along the Atlantic Ocean. The climate is equatorial: hot and humid. Relative humidity is high and rarely below 80%. Average monthly temperatures are around 27°C all year. Rainfall shows a significant seasonal variation due to the influence of migration from the Intertropical Convergence Zone (ZCIT). The average annual rainfall is about 3 meters. These high accumulations are mainly recorded during the rainy season, from December to June. During the dry season, from July to November, rainfall is low. There are large differences in annual rainfall accumulation from year to year. This phenomenon is governed by El Niño. During El Niño periods, French Guiana is subjected to precipitation deficits. On the other hand, La Niña leads to a higher annual rainfall accumulations (Moron *et al*., 1995; Aceituno, 1988).

**Figure 4:**
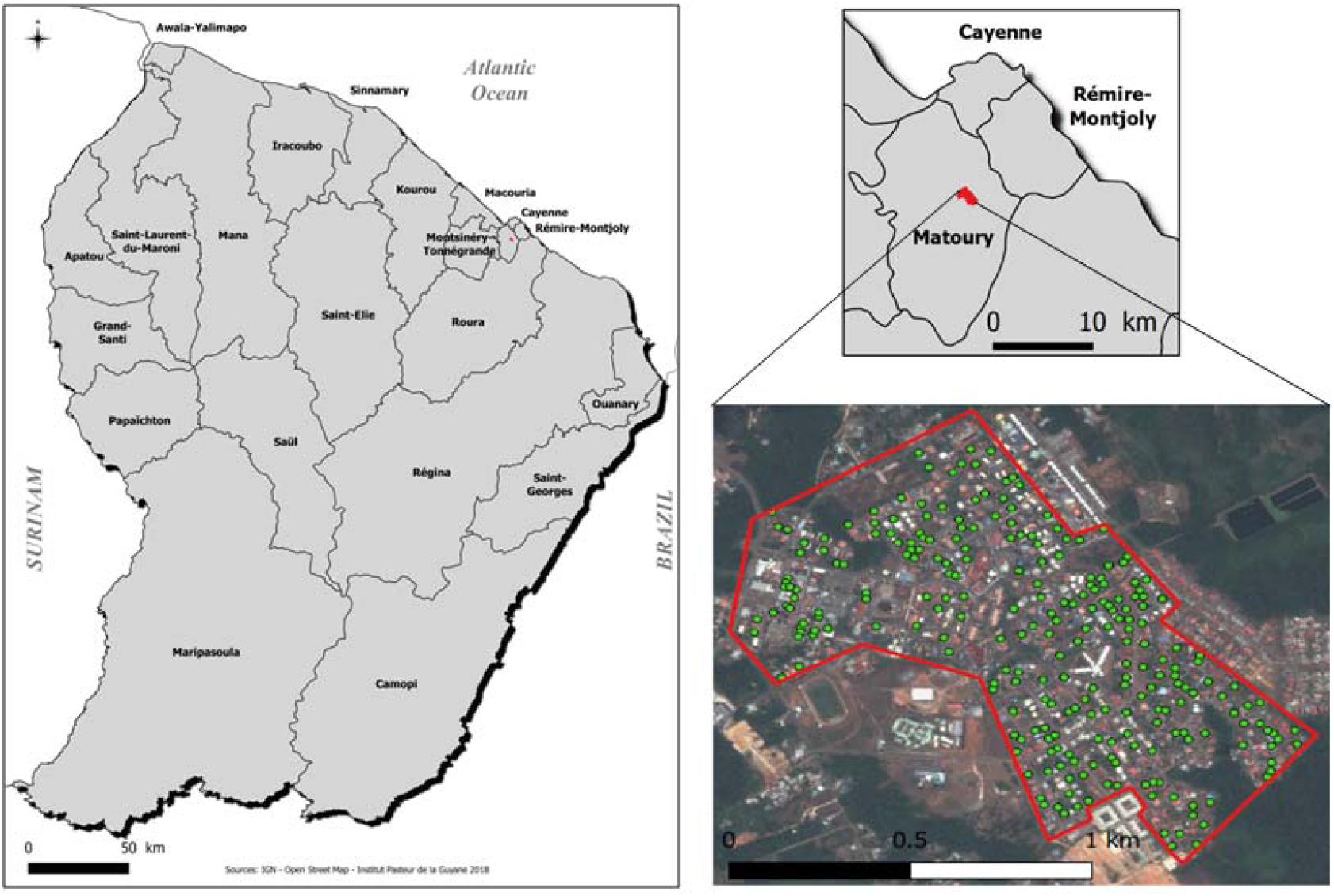
French Guiana, studied area and the center of Matoury.

Environmental conditions are more heterogeneous across the territory than meteorological conditions. Ninety percent of the territory is covered by the Amazon rainforest. The majority of the population lives along the coast where the three main municipalities are located: Cayenne and its surroundings, Kourou and Saint-Laurent-du-Maroni.

### Data collection

Entomological monitoring was carried out over a period of 18 months, from September 2011 to February 2013 to collect data in situ. This period included 2 months of dengue epidemics (Figure 5). Twenty houses per session (261 houses in total) were randomly sampled to (i) identify and count domestic and peridomestic containers and (ii) collect mosquitoes through the BG sentinel process. This trap releases artificial skin vapor to attract mosquitoes for blood meals and catch them in a bag. The trapping period was three days per house. Mosquitoes were then identified and counted by sex. These entomological surveys were conducted in the urban center of Matoury (Figure 4), a municipality neighboring Cayenne. With 217 inhabitants/km^2^ in 2012, this town has heterogeneous urbanization, with neighborhoods surrounded by dense forests. The center is made up of 875 easily accessible houses that are isolated from the rest of the town, which are representative of the urbanization of French Guiana.

**Figure 5:**
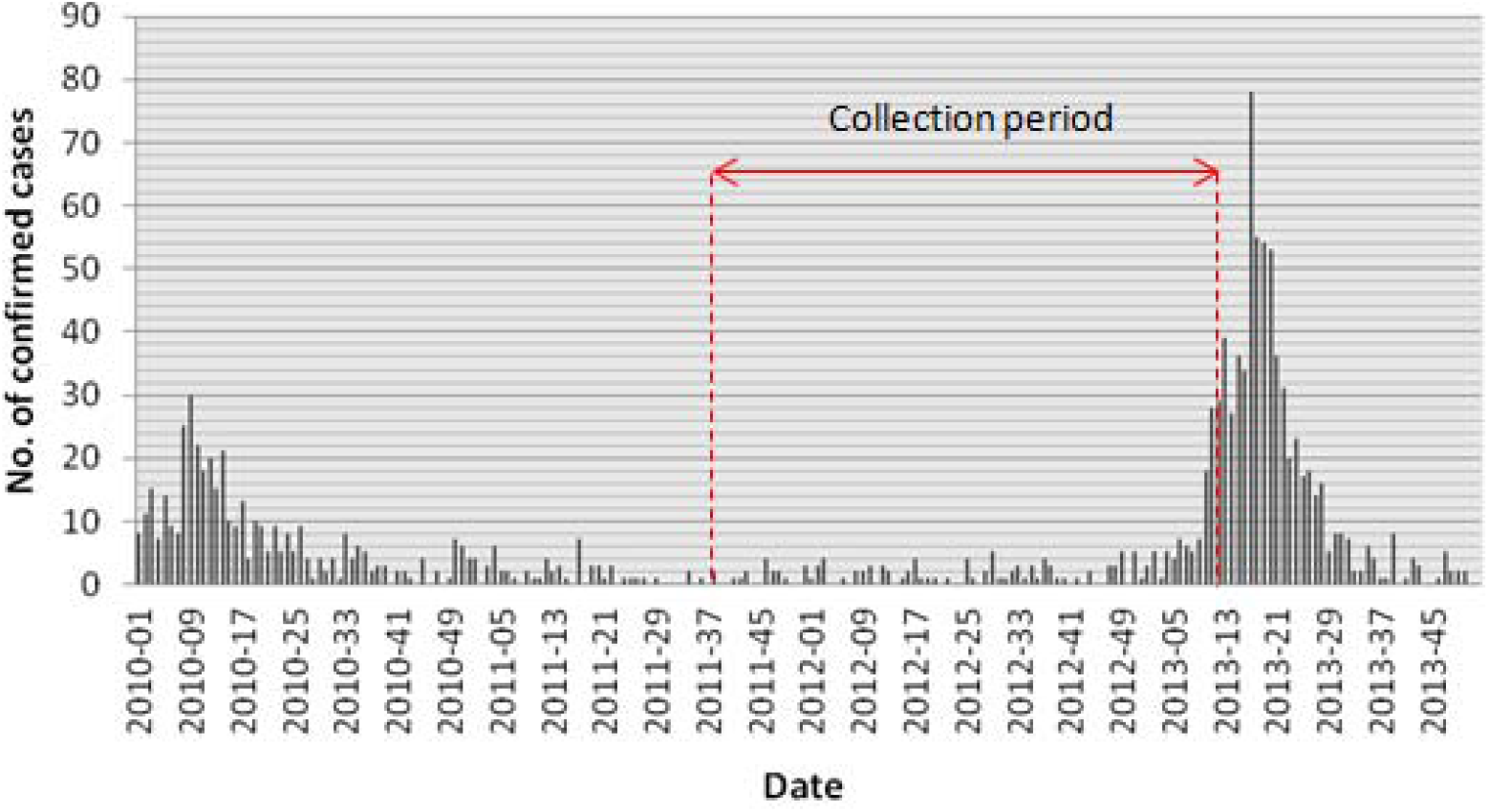
Number of confirmed cases collected by the regional system of epidemiological surveillance and entomological collection period in Matoury, French Guiana.

Daily meteorological data were obtained from the Météo France “Félix Eboué” station, based at the Matoury airport. For each trapping date, 243 variables were averaged or cumulated over different previous periods: 5, 10, 15, 20, 25 and 30 days. A lag effect was integrated with characteristics calculated over the last 5-10, 5-15, 5-20, 5-25 and 5-30 days. Satellite observations of rainfall (TRMM Tropical Rainfall Measuring Mission: the pixel closest to the area or the average of the surrounding pixels) were also available to increase the spatial information of rainfall data over the territory.

The Pleiades satellite constellation delivers images that combine high coverage, a high resolution (50 cm) and a high update rate: every point in the world is flown over every three to four days, with a resolution of 50 cm. Pairs of 4 optical Pleiades images covering the dry season and the beginning of the rainy season were acquired. They were acquired on 23/09/2012 and 10/12/2013 respectively. The images were projected in WGS 84, UTM Zone 22 N and geometrically corrected using the SRTM 90 m database produced by NASA. Three corrections were performed on the images: radiometric, atmospheric and geometric corrections. Image processing was performed using ENVI 5.1 and ENVI EX (Exelis Visual Information Solutions).

To calculate the different indices, the Orfeo Toolbox (OTB) was used. Each index highlights spectral characteristics related to the physical and biological characteristics of the field components. These indices are grouped into three categories: vegetations, soil and water indices. Fourteen indices were calculated. The indices taking into account atmospheric effects (e.g. GEMI) were calculated from the images before the atmospheric correction was applied.

For each image, a land cover map was produced containing the following classes: building, pool, asphalt, water, bare ground, light vegetation, green grass and tree. This map is the result of two classifications. The first is a supervised maximum likelihood pixel classification, performed in ENVI 5.1. For each class identified, a set of training polygons was digitized by an operator who interpreted the optical image. The spectral signature of each class was then constructed by the software. Each pixel was assigned to the class with the highest probability of being the correct pixel based on these spectral signatures. The kappa coefficient, which provides a measure of the accuracy of the classification, was calculated. This coefficient was 0.93 for each image, indicating good agreement between the resulting classes and the validation areas. In a second step, object-oriented classification was performed to improve the classification (e.g. to avoid class confusion). Object-oriented classification takes into account not only the spectral information of the pixels but also their spatial environment. Segmentation and merging of objects based on rule operations were carried out in the ENVI FX module of the ENVI software. Using confusion trees between the pixel and object classifications, it was possible to better classify objects such as blue or white roofs and swimming pools.

A geographic information system (GIS) was built in ArcGIS 10.2 to extract environmental variables at the experimental unit scale, *i*.*e*. at the house level. For each house identified by its centroid (extracted from the IGN cadastral data), various information was extracted from the immediate environment: the averages of all indices and the land use class zones were calculated. Each variable was calculated for buffer zones 10, 20, 30, 40, 50, 100, 200, 300, 400 and 500 m around the house. For each house, the distance to the first pixel of each class was calculated, as well as the area of each land use class around the experimental unit.

### Statistical and spatial analyses

A large heterogeneous database was constructed to centralize the entomological data with the 747 geographic indicators and 195 meteorological indicators. Univariate logistic regressions were used to model the probability of the presence of larvae testing each of the 942 meteorological or environmental as explanatory variables. All variables with a p-value<0.25 were retained in the initial set of BRT variables. A total of 317 explanatory variables were retained by this method.

The analysis of the presence/absence of larvae was based on the boosted regression tree (BRT) method (Friedman *et al*., 2000) and fitted to a “Bernoulli” distribution. This is a “machine learning” method for characterizing the shape of the relationship between the variable of interest and a large number of explanatory variables. BRTs are able to work with a large number of complex variables with interactions or nonlinearities (Elith *et al*., 2008). This method has been used in various fields of study, including disease modeling (Elith *et al*., 2008, Stevens and Pfeiffer, 2011 and Cheong *et al*., 2014). BRTs combine the strengths of two algorithms: regression trees (models that link predictors to responses through a recursive sequence of binary separations) and boosting (which allows the combination of several simple models to improve predictive performance). The aim of BRTs is to combine several simple models that together will provide a good prediction. The model is built by adding successive regression trees in an iterative way, with each addition the performance of the model is measured by a cross-validation. The model building process is optimal when the learning rate (lr) is slower, also called the decay rate. The complexity of the tree (tc) determines the maximum degree of interaction between the predictors with respect to the response. The more levels of interaction there are, the higher the tc value is. After several trials, we chose a tc of 5 with a lr of 0.001. The initial number of trees (nt) was 50, with subsequent trees being added by iteration. The part of observations not used to build the model but used to perform the cross-validation (bag fraction) was set to 0.5. The measure of the strength of the association between each explanatory variable and the outcome was given by a percentage of influence; the total influence was 100% (Friedman, 2001). A higher percentage of a variable indicates a higher relative importance of that variable on the response. The ROC curve is the probability that the classifier scores higher than a randomly drawn positive sample. If the AUC is 1, there is no prediction error. An AUC = 0.5 means that the predictions are no better than random prediction. We applied a BRT using R version 3.0.2 (R Development Core Team, 2018), the “dismo” package version 0.8-17 (Elith *et al*., 2008; Hijmans *et al*., 2020) and the “gbm” package version 2.1 (Ridgeway, 2020).

The model created was extrapolated to the municipalities of Cayenne, Rémire-Montjoly and Matoury during part of the last dengue epidemic period in order to compare the location of reported cases to the hot-spots identified by the model. November 2019 to August 2020. Data corresponding to the variables identified in the model were collected. Two meteorological stations allowed us to obtain a meteorological dataset without any missing data. One of the stations was the same as that used to create our model. The second station that provided us with weather data was located in Rémire-Montjoly (Figure 4). We used a Pleiades image taken on 20 December 2017. This was the most recent image with a minimal clouds cover.

In addition to creating entomological risk maps for the three municipalities, the extrapolated data allowed the assessment of correlations between entomological variables (model predictions) and epidemiological variables (geolocated biologically confirmed cases from November 2019 to August 2020). To determine whether the areas of the three municipalities were more affected by the predictions and/or by the number of biologically confirmed cases, the data were averaged according to different geographical entities of different sizes: 500 × 500 m and 1000 × 1000 m grids, as well as by IRIS (homogenous geographical and demographic micro neighborhoods). The different grid sizes studied allow us to determine the most appropriate scale for studying the results of the model.

For this purpose, correlation matrices at these different scales and between the entomological and epidemiological variables were created. These correlations of variables were also obtained using choropleth maps that represent the data by discretized ranges of values.

## Data Availability

Epidemiological aggregated data belonging to French Public Health Agency (http://www.santepubliquefrance.fr/ <http://www.santepubliquefrance.fr/>) can be obtained contacting the scientific coordinator of Guyane Regional Epidemiology Unit of the French Public Health Agency. All interested researchers can access the data by the same means the authors accessed them. Clinical data will not be made available according to the French CNIL recommandations (Commission Nationale Informatique et Libertes) that require specific authorizations to transfer data from one center to another. Requests for data transfer can be made to: CNIL, 8 rue Vivienne, CS 30223, 75083 Paris cedex 02, France. Tel: +33 (0)1.53.73.22.22; Fax: +33 (0)1.53.73.22.00.

## Acknowledgements

This study was supported by the Centre National d’Etudes Spatiales-Terre solide, Océan, Surfaces Continentales, Atmosphère fund (grant number CNES-TOSCA-4800000720). CF acknowledges the European Regional Development Fund under EPI-ARBO grant agreement GY0008695.

## Ethics statement

Laboratory surveillance data collection issued from the national surveillance system and was approved by the Advisory Committee on Information Processing in Material research in the Field of Health (N°07.148) and was authorized by regulatory authorities (CNIL-N°1213498).

